# COVID-19 pandemic and health worker stress: The mediating effect of emotional regulation

**DOI:** 10.1101/2020.06.19.20135574

**Authors:** Zoilo Emilio García-Batista, Kiero Guerra-Peña, Vahid Nouri Kandany, Maria Isabel Marte, Luis Eduardo Garrido, Luisa Marilia Cantisano-Guzmán, Luciana Moretti, Leonardo A. Medrano

## Abstract

**Background/Introduction:** Psychological and physical well-being of health personnel has been significantly affected by COVID-19. Work overload and continuous exposure to positive COVID-19 cases have caused them fatigue, stress, anxiety, insomnia and other detriments. This research aims: 1) to analyze whether the use of cognitive reevaluation and emotional suppression strategies decreases and increases, respectively, stress levels of health personnel; 2) to quantify the impact of contact with patients with COVID-19 on stress’s level of medical staff.

**Method:** Emotion regulation strategies and stress level were evaluated in 155 Dominican physicians by means of psychological tests with adequate levels of reliability. In addition, a questionnaire created by the researchers quantified the impact that contact with those infected had on their stress levels.

**Results:** Contact with patients with COVID-19 predicts increased use of emotion suppression strategies, although is not associated with the use of cognitive reevaluation. These findings lead to an even greater increase in stress on health care providers.

**Conclusions:** Contextual contingencies demand immediate responses and may not allow health personnel to use cognitive re-evaluation strategies, leaning more towards emotion suppression. However, findings regarding high levels of stress require the implementation of intervention programs focused on the promotion of more functional emotion regulation strategies. Such programs may reduce current stress and prevent post-traumatic symptoms.

## Introduction

Coronavirus pandemic (COVID-19) continues to spread internationally, putting increased pressure on health care workers. The need to provide immediate responses and the volume of people infected generate an overload of work that increases levels of fatigue and stress [1]. In addition, the risks of exposure, concern about infecting loved ones, self-isolation measures and family-work conflict are factors that all together increase the likelihood of emotional disorders and problems associated with chronic stress [2-3].

Several studies indicate an increase in the prevalence of mental health symptoms among health workers who treat patients with COVID-19. A survey of 1257 physicians and nurses indicated that 50.4%, 44.6%, 34.0%, and 71.5% had symptoms of depression, anxiety, insomnia, and distress, respectively [4]. In a previous study during the acute SARS-Cov2 outbreak [2], 89% of health workers reported psychological symptoms. Sources of distress may include feelings of vulnerability or loss of control and concerns about one’s health, the spread of the virus, the health of family and others, changes in work, and isolation.

This situation is even more complex for health professionals in developing countries [5], as is the case in the Dominican Republic. The lack of sufficient resources for patients’ treatment and health worker protection [6] increases the overload of health workers and the risk of experiencing stress-related problems.

Faced with this scenario, various agencies have highlighted the need to address the psychological safety of health workers [7]. As the Pan American Health Organization [8] points out, attending to the mental health and psychosocial well-being of health workers is as important as taking care of their physical health. However, psychological factors that could moderate the levels of psychological stress during the course of the pandemic have not yet been studied empirically.

Within this framework, emotion regulation (ER) strategies play a significant role. In the last decade, there has been increasing interest in exploring how people manage or regulate their emotions through specific strategies. The model of emotion regulation process is one of the most influential theoretical proposals to outline the mechanisms by which people modulate their emotions. Within this model, two well-defined ER strategies have been empirically explored: Cognitive reevaluation (CR), a cognitive strategy that involves redefining a potentially emotive situation in such a way as to change its emotional impact; and expressive suppression (ES), a form of response modulation that involves inhibiting the expressive behavior of the emotion in progress [9].

These strategies have been differentially associated with psychological and health adjustment variables, converging to the negative effects of suppression and the positive effects of reevaluation [10]. Thus, CR has been positively correlated with self-esteem, optimism, personal growth and purpose in life, while inverse correlations have been reported with the negative effects, stress and depression [9, 11]. On the other hand, SA increases physiological activity and has negative effects on memory, and has been positively associated with negative affect, anxiety and depression [12, 13].

Overall, previous findings lead to the assumption that health personnel who make adequate use of ER strategies will have lower levels of perceived stress. Conversely, those professionals with greater difficulties in regulating their emotions will present greater symptoms associated with stress [14-16].

It should be noted that analyzing the factors involved in the appropriate stress regulation is not only a relevant issue for the psychological well-being of health-care workers, but also for the patients themselves. Inadequate stress regulation can diminish the empathy that health personnel may have towards patients, reduce impulse control, increase aggression and, in general, affect the quality of their services. In addition, high levels of stress can lead to health personnel making attentional mistakes, such as medication failure or mistakes in the implementation of patient care techniques [16-18].

Depending on the importance of identifying protective factors of stress in health-care workers, the present study aims to analyze whether ER strategies have a mediating role on perceived stress of health-care workers. More specifically, it is hypothesized that the use of cognitive reevaluation strategies decreases stress levels of health-care workers and that emotion suppression strategies increase stress levels. In addition, it is intended to quantify the impact of contact with patients with COVID-19 (number of patients and hours spent) and of the team’s perceived safety on medical staff’s stress.

## Methods

This research had the revision and approval of the National Council of Bioethics in Health/ Consejo Nacional de Bioética en Salud (CONABIOS) of the Dominican Republic. The protocol registration number in CONABIOS was −005-2019.

### Participants

The sample was composed of 155 physicians (67.9% women and 32.1% men) from the Dominican Republic, ranging in age from 23 to 66 (mean age = 34.89; SD = 9.26).

### Instruments

#### Perceived Stress Scale (PSS-14)

This instrument was designed by Cohen et al. [19] to measure the degree to which life situations are perceived as stressful during the last month. Its approximate application time is 8-10 min, and it is made up of 14 direct and indirect items. It uses a Likert-type response format of 5 alternatives, with a range from 0 (never) to 4 (very often), inverting the score on items 4, 5, 6, 7, 9, 10 and 13. The scale scores from 0 to 56; higher scores indicate greater perceived stress. This scale has demonstrated in several populations to have consistent psychometric properties for the measurement of stress [20].

#### Emotion Regulation Questionnaire (ERQ) [10]

This instrument is designed to evaluate emotion regulation strategies by means of 10 items, in detail, 6 items that assess cognitive reevaluation and 4 items that represent the suppression of emotional expression. These are evaluated by a Likert-type scale with 7 response options ranging from 1 (totally disagree), 2 (disagree), 3 (slightly disagree), 4 (neither agree nor disagree), 5 (slightly agree), 6 (agree), to 7 (totally agree) [21-23].

On the other hand, a questionnaire was designed with questions regarding the number of hours treating people with COVID-19 and the amount of patients treated, questions about the degree of perceived safety in the equipment used, their rank within the health institution and type of health-care center.

## Statistical Analyses

### Modelling specifications

A structural equation model (SEM) was estimated in order to assess the mediating effects of ER on the impact of COVID-19 contact and equipment safety on perceived stress of health-care workers. Because the model contained a mixture of continuous and categorical variables, the weighted minimum squares with mean- and variance-adjusted standard errors (WLSMV) estimator was employed, which is widely recommended for models that include ordinal-categorical variables [24]. As the underlying structure of the scores from the Emotion Regulation Questionnaire items is composed of two factors, they were estimated using an exploratory structural equations model (ESEM) [25], with factors rotated using the Geomin algorithm [26]. In general, psychological measures are fallible or impure indicators of their underlying trait, and as such, the factor structures containing them are more accurately estimated with unrestricted models that allow the items to load freely on different factors [27, 28].

The significance and confidence intervals of the indirect effects was evaluated using bootstrapping, which has demonstrated optimal functioning [29]. A total of 50,000 random samples with replacement were generated from the empirical data, and the 95% confidence intervals were constructed by taking the values corresponding to 2.5 and 97.5 percentiles of the parameter estimate distribution. In order to combine bootstrapping with ESEM factors, the ESEM within CFA method was employed [28]. Also, to evaluate the size of the mediation effects, Cohen’s [30] benchmarks of .01 for small, .09 for medium, and .25 for large effects were used for the completely standardized indirect effects (*ab*_*cs*_) [31].

Wording effects resulting from the Perceived Stress Scale have been balanced, with half the items reversed coded, were modeled using random intercept item factor analysis (RIIFA) [32]. The RIIFA model adds a wording method factor where the pro-trait items have loadings of +1 and the *recoded* reversed items have loadings of −1. Thus, it posits an artifactual relationship between the groups of items that contrasts with the substantive factor, where all the items are expected to have loadings of the same sign. Additionally, the wording factor was specified to be uncorrelated with the substantive factor in order to ensure identification. The RIIFA model has performed well in accounting for wording variance arising from the responses to scales that combine items of opposite polarity [33, 34].

### Fit criteria

The fit of the SEM model was assessed with four complimentary indices: the comparative fit index (CFI), the Tucker-Lewis index (TLI), the root mean square error of approximation (RMSEA), and the standardized root mean square residual (SRMR). Values of CFI/TLI greater than or equal to .90 and .95 have been suggested that reflect acceptable and excellent fits to the data, while values of RMSEA less than .08 and .05 may indicate reasonable and close fits to the data, respectively [35-37]. In the case of SRMR, a value less or equal to .08 has been found to indicate a good fit to the data [35, 38]. It should be noted that because the values of these fit indices are also affected by incidental parameters not related to the size of the misfit [39-41], they should not be considered golden rules, and must be interpreted with caution [36, 42].

### Reliability analyses

The internal consistency reliability of the psychological scale scores was evaluated with Green and Yang’s [43] categorical omega coefficient. Categorical omega takes into account the ordinal nature of the data to estimate the reliability of the observed scores, and as such, it is recommended for Likert-type item scores [44, 45]. In order to provide common reference points with the previous literature, Cronbach’s [46] alpha with the items treated as continuous was also computed and reported. Additionally, the reliability of the scores for COVID-19 contact, which were derived from two continuous measures, was estimated using alpha based on the standardized scores [47]. For all coefficients 95% confidence intervals were computed across 1,000 bootstrap samples using the bias-corrected and accelerated approach [48].

### Missing data handling

Missing data for the variables included in the SEM model was very small, with only a 0.3% total missing value rate. None of the items from the Perceived Stress Scale had missing values, while two items from the Emotion Regulation Questionnaire had one missing value each (0.6% rate). Neither age, sex, or the two COVID-19 contact items had missing values. Finally, the variable measuring the perceived safety provided by the protective equipment had 7.7% cells with missing values. According to Little’s [49] MCAR test the data were missing completely at random (χ^2^ = 105.11, *df* = 84, *p* = .059). Due to the very small amount of missingness and the MCAR mechanism, the missing data was handled using pairwise deletion [50].

### Analysis software

Data handling, descriptive statistics, and Little’s MCAR test were computed using the IBM SPSS software version 25. Sample correlations and the SEM model were estimated with the *Mplus* program version 8.3. Internal consistency reliability with the categorical omega and alpha coefficients was estimated with the *ci*.*reliability* function contained in the *MBESS* package (version 4.6.0) [51].

## Results

Health-care professionals that participated in this study worked an average of 4.49 hours daily (SD = 4.17) with COVID-19 patients and had daily contact with an average of 2.46 people (SD = 3.81) infected with the virus. Regarding the perceived safety provided by their protective equipment, the mean scores were 3.35 (SD = 2.66) on the 1-10 response scale. Additionally, the mean scores were 1.78 (SD = 0.64) across the perceived stress items (0-4 scale), 3.45 (SD = 0.79) across the cognitive reevaluation items (1-5 scale), and 2.93 (SD = 1.05) across the emotion suppression items (1-5 scale). The sample correlations between the observed variables included in the SEM model are presented in Table A1 in the Appendix.

According to the categorical omega reliability coefficient, all the scales had adequate internal consistency reliability. In the case of the perceived stress scores, the reliability estimate was .928 (95% CI = .894, .943). For the cognitive reevaluation scale, the reliability estimate was .723 (95% CI = .595, .794), while for emotional suppression it was .762 (95% CI = .663, .822). In order to provide a common reference with previous studies, the (suboptimal) alpha estimates for these scales’ scores were: .898 (95% CI = .873, .919) for perceived stress, .682 (95% CI = .582, .760) for cognitive reevaluation and .749 (95% CI = .665, .815) for emotion suppression. Finally, the COVID-19 composite score had a reliability of .639 (95% CI = .399, .743) according to the alpha coefficient.

Figure 1 depicts a simplified version of the estimated SEM model that assessed the mediating effects of emotion regulation on the impact of COVID-19 contact and equipment safety on the perceived stress of medical personnel. In order to statistically control age and sex in the SEM model, all the latent variables, except the wording factor as well as the equipment perceived safety item, were regressed on them. Also, because age and sex were exogenous variables, they were allowed to correlate. As typical, the residuals from endogenous variables that shared the same predictors were allowed to correlate.

**Figure 1.**
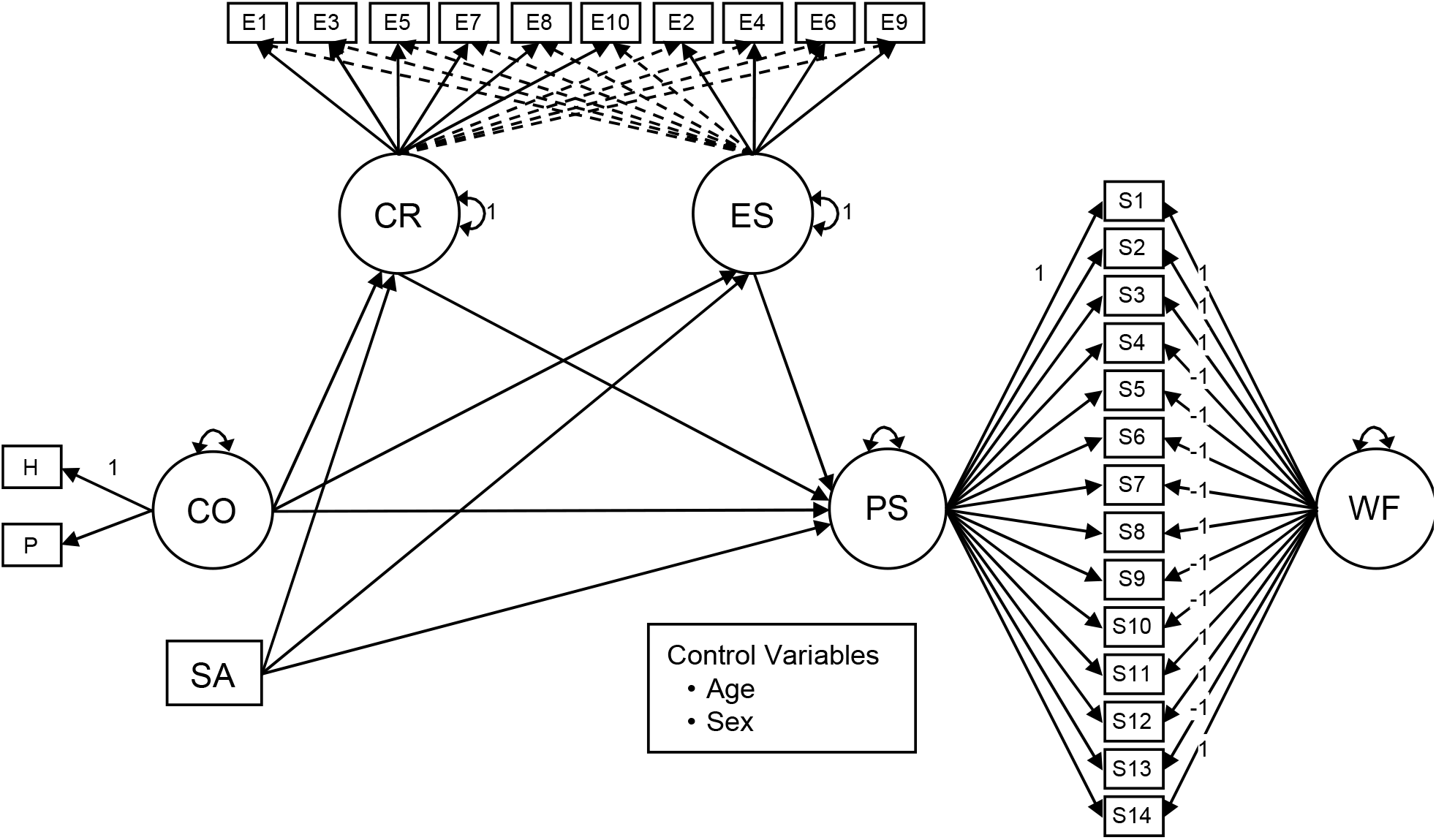
SEM model evaluating the mediating effects of emotional regulation on the impact of COVID-19 contact and equipment safety on the perceived stress of medical personnel. *Note*. CO = contact with COVID-19 patients; SA = perceived safety provided by the protective equipment; PS = perceived stress; CR = cognitive regulation; ES = emotional suppression; WF = wording factor; H = daily number of hours treating COVID-19 patients; P = daily number of COVID-19 patients treated; S1-S14 = Perceived Stress Scale items; E1-E10 = Emotional Regulation Questionnaire items. Squares represent observed variables. Circles represent latent variables. Full unidirectional arrows linking circles and rectangles represent the target factor loadings. Dotted unidirectional arrows linking circles and rectangles represent the cross-loadings. Bidirectional arrows connecting a single circle represent the factor variances. For simplicity, the control variables do not appear represented in the model, as well as the item uniquenesses, the factor uniquenesses, and the residual correlations between variables that share the same predictors.

According to the different indices evaluated, the fit of the estimated SEM model was good: χ^2^_350_ = .515.43 (*p* < .001), CFI = .944, TLI = .935, RMSEA = .055 (90% CI = .045, .065), and SRMR = .063. The item factor loadings derived from the SEM model are presented in Table 1. In general, the factors had adequately sized factor loadings. The mean factor loadings were .68 for the perceived stress factor, .58 for cognitive reevaluation, .63 for emotion suppression, and .69 for COVID-19 contact. As expected, the items from the Emotion Regulation Questionnaire produced several significant cross-loadings of considerable magnitude, supporting the use of ESEM modeling for the two factors derived from this instrument. On the other hand, the loadings on the wording method factor were significant and of moderate magnitude (.233), revealing the presence of wording variance in the perceived stress item scores.

**Table 1.** Item factor loadings for the estimated SEM model

| Item | Factors |  |  |  |  |
| --- | --- | --- | --- | --- | --- |
|  | CO | PS | WF | CR | ES |
| H | .594** |  |  |  |  |
| P | .791** |  |  |  |  |
| S1 |  | .715** | .233** |  |  |
| S2 |  | .792** | .233** |  |  |
| S3 |  | .816** | .233** |  |  |
| S4 |  | .718** | -.233** |  |  |
| S5 |  | .752** | -.233** |  |  |
| S6 |  | .698** | -.233** |  |  |
| S7 |  | .673** | -.233** |  |  |
| S8 |  | .567** | .233** |  |  |
| S9 |  | .802** | -.233** |  |  |
| S10 |  | .761** | -.233** |  |  |
| S11 |  | .734** | .233** |  |  |
| S12 |  | .255** | .233** |  |  |
| S13 |  | .429** | .233** |  |  |
| S14 |  | .754** | .233** |  |  |
| E1 |  |  |  | .577** | -.007 |
| E3 |  |  |  | .413** | .088 |
| E5 |  |  |  | .535** | -.499** |
| E7 |  |  |  | .616** | .255* |
| E8 |  |  |  | .646** | .271* |
| E10 |  |  |  | .685** | -.079 |
| E2 |  |  |  | -.062 | .792** |
| E4 |  |  |  | .008 | .595** |
| E6 |  |  |  | .168 | .692** |
| E9 |  |  |  | .290** | .449** |

Note. CO = contact with COVID-19 patients; PS = perceived stress; WF = wording factor; CR = cognitive regulation; ES = emotional suppression; H = daily number of hours treating COVID-19 patients; P = daily number of COVID-19 patients treated; S1-S14 = Perceived Stress Scale items; E1-E10 = Emotional Regulation Questionnaire items.
\* $p < .05$ , \*\* $p < .01$ .

The standardized direct and indirect regression weights (β), residual correlations, and correlations from the estimated SEM model are shown in Table 2. The main findings from the results included in the table are: *first*, contact with COVID-19 patients increased emotion suppression (β = .363, *p* = .002) of the medical personnel, but not their cognitive reevaluation (β = .124, *p* = .384). *Second*, cognitive reevaluation decreased the perceived stress (β = −.425, *p* < .001), while emotion suppression increased it (β = .645, *p* < .001). *Third*, emotion suppression mediated the effects of COVID-19 contact with a near large effect size (β = .234, *p* < .01), but cognitive reevaluation was not a significant mediator (β = −.053, *p* > .05). *Fourth*, contact with COVID-19 patients did not have a direct effect on perceived stress (β = .122, *p* = .183). *Fourth*, as the perceived safety provided by the protective equipment did not affect emotion suppression (β = −.133, *p* = .209), cognitive reevaluation (β = .041, *p* = .653), or perceived stress (β = .083, *p* = .302) of the personnel, neither cognitive reevaluation (β = .-.018, *p* > .05) nor emotion suppression (β = .-.086, *p* > .05) were significant mediators in relation to this variable. *Sixth*, older workers had less contact with COVID-19 patients (β = −.238, *p* = .025), and reported more cognitive reevaluation (β = .269, *p* = .003). *Seventh*, males reported less cognitive reevaluation (β = −.510, *p* < .001) and less emotion suppression (β = −.235, *p* = .023) than females. *Finally*, the proportion of variance explained of the mediators and dependent latent variables were: .244 (*p* < .001) for cognitive reevaluation, .243 (*p* = .002) for emotion suppression and .578 (*p* < .001) for perceived stress.

**Table 2.**
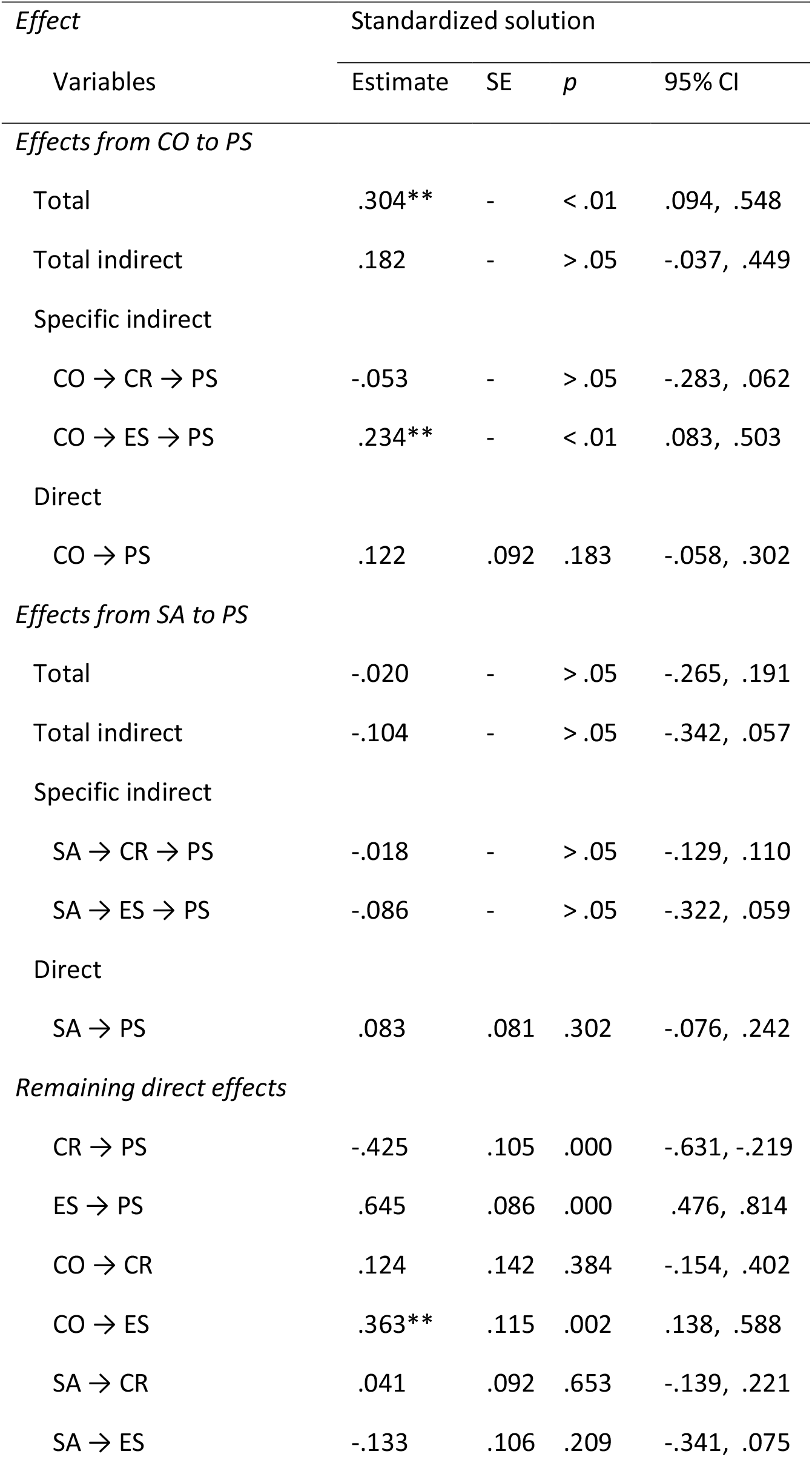
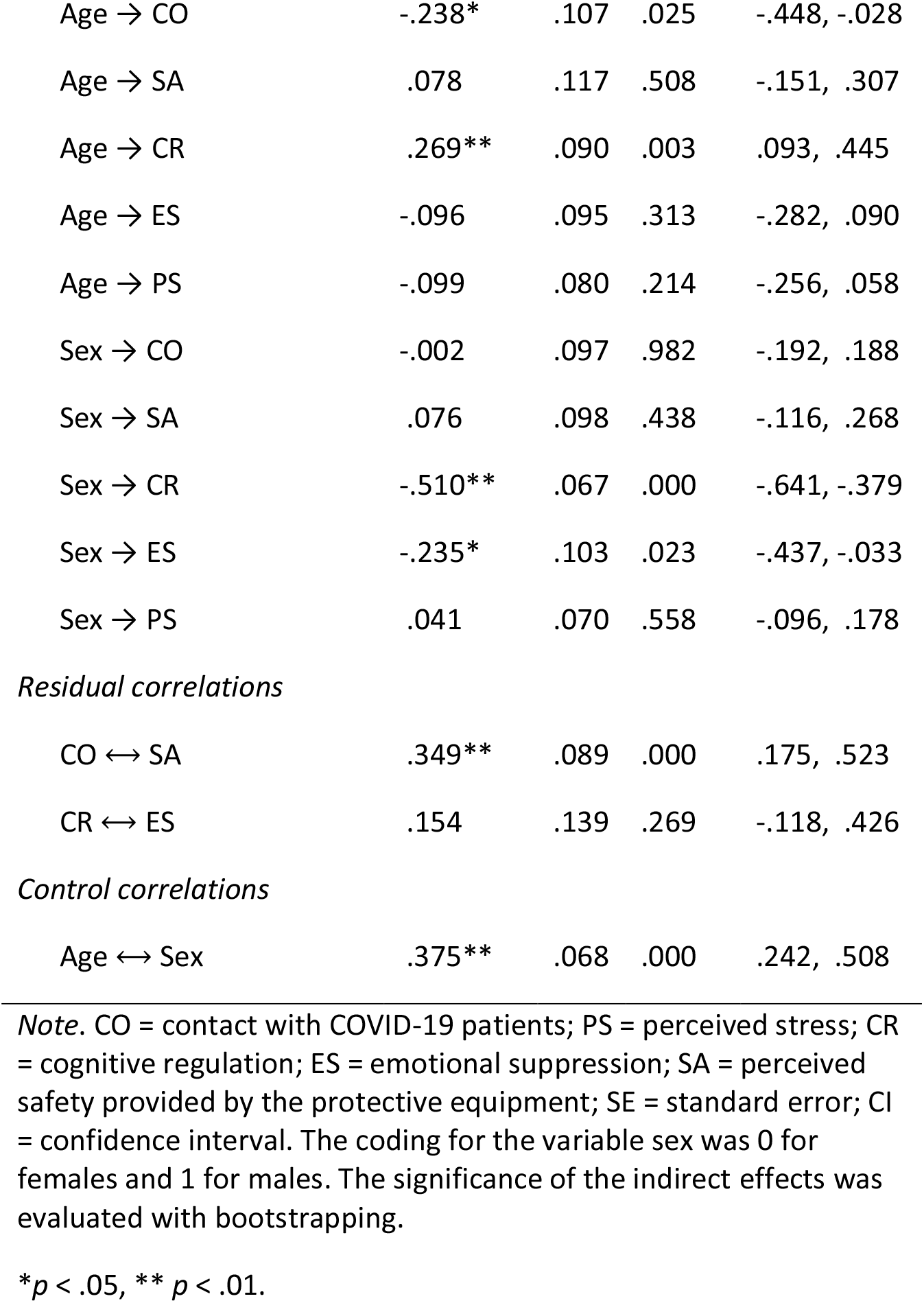
Regressions weights and correlations from the estimated SEM model

| *Effect*<br>Variables | Standardized solution |  |  |  |
| --- | --- | --- | --- | --- |
|  | Estimate | SE | *p* | 95% CI |
| *Effects from CO to PS* |  |  |  |  |
| Total | .304** | - | < .01 | .094, .548 |
| Total indirect | .182 | - | > .05 | -.037, .449 |
| Specific indirect |  |  |  |  |
| CO → CR → PS | -.053 | - | > .05 | -.283, .062 |
| CO → ES → PS | .234** | - | < .01 | .083, .503 |
| Direct |  |  |  |  |
| CO → PS | .122 | .092 | .183 | -.058, .302 |
| *Effects from SA to PS* |  |  |  |  |
| Total | -.020 | - | > .05 | -.265, .191 |
| Total indirect | -.104 | - | > .05 | -.342, .057 |
| Specific indirect |  |  |  |  |
| SA → CR → PS | -.018 | - | > .05 | -.129, .110 |
| SA → ES → PS | -.086 | - | > .05 | -.322, .059 |
| Direct |  |  |  |  |
| SA → PS | .083 | .081 | .302 | -.076, .242 |
| *Remaining direct effects* |  |  |  |  |
| CR → PS | -.425 | .105 | .000 | -.631, -.219 |
| ES → PS | .645 | .086 | .000 | .476, .814 |
| CO → CR | .124 | .142 | .384 | -.154, .402 |
| CO → ES | .363** | .115 | .002 | .138, .588 |
| SA → CR | .041 | .092 | .653 | -.139, .221 |
| SA → ES | -.133 | .106 | .209 | -.341, .075 |
| Age → CO | -.238* | .107 | .025 | -.448, -.028 |
| Age → SA | .078 | .117 | .508 | -.151, .307 |
| Age → CR | .269** | .090 | .003 | .093, .445 |
| Age → ES | -.096 | .095 | .313 | -.282, .090 |
| Age → PS | -.099 | .080 | .214 | -.256, .058 |
| Sex → CO | -.002 | .097 | .982 | -.192, .188 |
| Sex → SA | .076 | .098 | .438 | -.116, .268 |
| Sex → CR | -.510** | .067 | .000 | -.641, -.379 |
| Sex → ES | -.235* | .103 | .023 | -.437, -.033 |
| Sex → PS | .041 | .070 | .558 | -.096, .178 |
| *Residual correlations* |  |  |  |  |
| CO ↔ SA | .349** | .089 | .000 | .175, .523 |
| CR ↔ ES | .154 | .139 | .269 | -.118, .426 |
| *Control correlations* |  |  |  |  |
| Age ↔ Sex | .375** | .068 | .000 | .242, .508 |

*Note.* CO = contact with COVID-19 patients; PS = perceived stress; CR = cognitive regulation; ES = emotional suppression; SA = perceived safety provided by the protective equipment; SE = standard error; CI = confidence interval. The coding for the variable sex was 0 for females and 1 for males. The significance of the indirect effects was evaluated with bootstrapping.
\* $p < .05$ , \*\* $p < .01$ .

## Discussion

COVID-19 has revolutionized the world and had a great impact on the physical and mental health of millions of people [4]. Concern about infection or transmission to a family member, social isolation, and economic impact have led to an increase in the prevalence of stress-related problems in the general population [52]. However, the impact of this pandemic on stress is especially critical for health-care workers.

The consequences of high and chronic stress are multiple. First, it affects the mental health of workers, as suffering from occupational stress doubles the probability of developing a mental disorder [53, 54], and predicts the development of anxious and depressive clinical symptoms [55]. Secondly, it is associated with the development of physical diseases, such as cardiovascular problems [14-16, 56]. Finally, stress can diminish the empathy of health-care professionals towards patients, reduce their impulse control and affect the quality of their services [16-18].

This situation is even more critical in countries with fewer health resources [5], such as the Dominican Republic. The lack of sufficient resources for the treatment of patients and for the protection of health-care workers [6] increases the overload of them and the risk of experiencing problems associated with stress. On the other hand, it leads to the need for health-care professionals to make ethically and morally difficult decisions about those who receive these scarce resources. Their decisions can mean life or death for many. This can cause chronic stress, moral damage, and feelings of guilt [57].

Within this context, the identification of protective psychological factors that allow health-care professionals to reduce their stress levels and protect their mental health is critical. The results obtained in this study support the adjustment of a mediational model, where emotion regulation (ER) strategies play an important role on perceived stress levels.

ER refers to a set of processes aimed to modulate the emotional state in order to respond to a series of external demands in an appropriate way [9]. Results indicate that when exposed to contact with patients with COVID-19, health-care workers tend to use predominantly strategies of emotion suppression. In this regard, it should be noted that they probably have no other alternative, since faced with the need to give an immediate response; doctors make a deliberate effort to limit emotion expression behaviors. Unfortunately, as results indicate, using this type of strategy increases stress levels.

In addition, previous research with refugees or people who were exposed to traumatic situations indicates that the use of emotion suppression strategies predicts the development of post-traumatic symptoms and increases the likelihood of developing mental problems in the future [58]. Furthermore, some studies suggest that emotion suppression is an aggravating factor in the effects of traumatic experience [59].

It is likely that environmental contingencies will not allow health-care professionals to make use of cognitive reevaluation strategies. As results indicate, contact with patients with COVID-19 predicts increased use of emotion suppression strategies, but is not associated with the use of cognitive reevaluation, which is shown to be inversely associated with stress levels.

These findings allow us to affirm that health-care workers are not only exposed to strong stressors, but that these environmental contingencies do not favor the deployment of more functional strategies of emotional regulation either.

For this reason, it is important that health-care workers receive support and containment through intervention programs focused on promoting more functional ER strategies [60, 61]. The aim would not be to avoid the use of emotion suppression, as this is probably the most appropriate strategy for dealing with these situational contingencies. Rather, the goal should be to promote a flexible use of emotion regulation strategies, which decreases stress levels and the likelihood of developing post-traumatic symptoms.

Based on all the above findings, it is imperative to develop measures and programs aimed at improving the mental health of health-care workers. This should be done as soon as possible, since inadequate emotion regulation not only puts at risk the psychological well-being of health-care professionals, but also patients’ health. When health-care workers are under great stress, they can make potentially fatal treatment failures [16-18]. The need for an immediate response is even greater when one considers that stress and emotional instability may result in lost workdays that would further limit the human resources that currently assist patients with COVID-19, making them a potential danger if a second wave of infection occurs.

## Data Availability

Data is available

## Acknowledgments

This research was supported by the National Fund for Innovation and Scientific and Technological Development (FONDOCYT) of the Dominican Republic, for this reason we thank you.

---

**Table A1.**
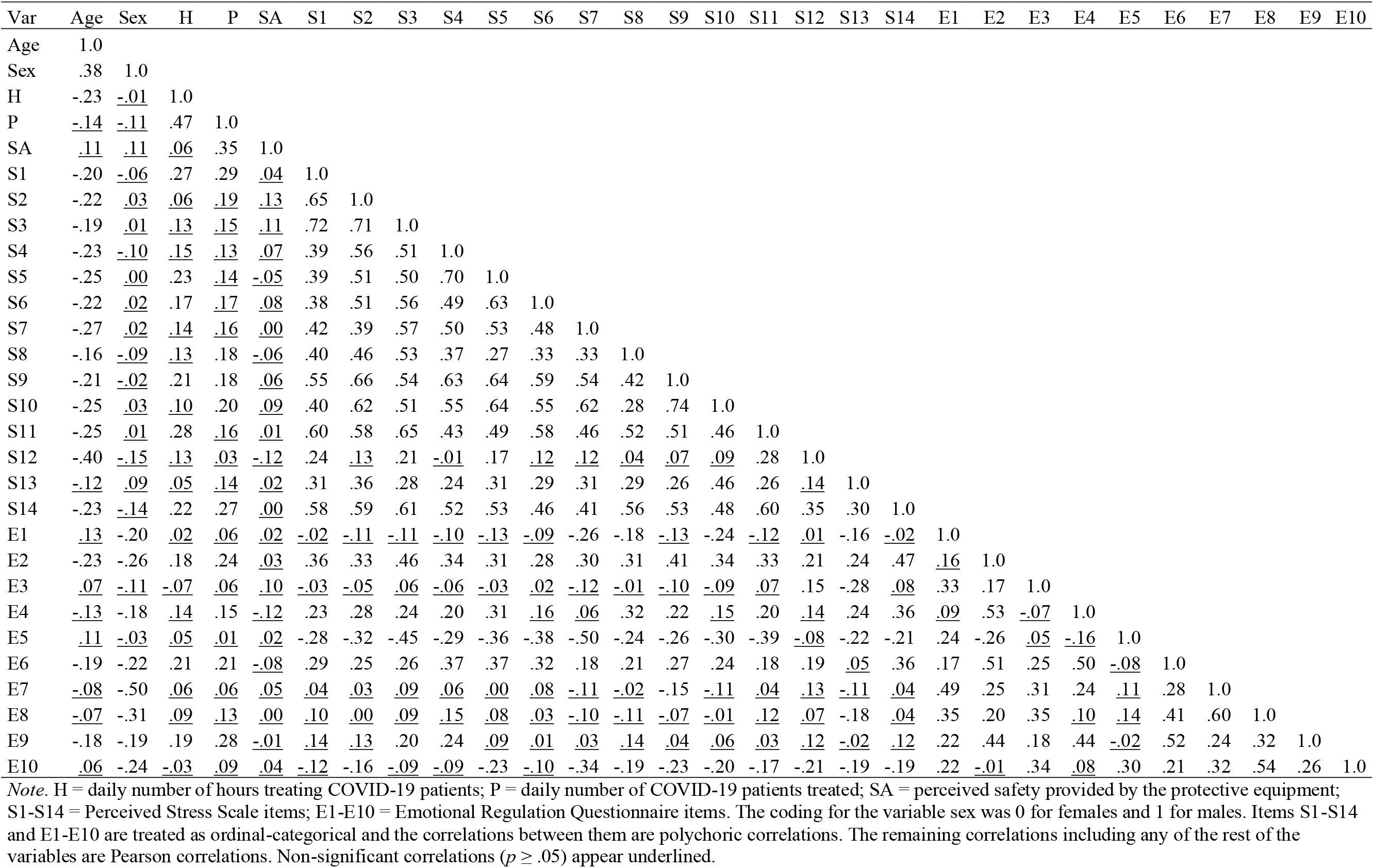
Sample correlations between the variables included in the SEM mediation model

| Var | Age | Sex | H | P | SA | S1 | S2 | S3 | S4 | S5 | S6 | S7 | S8 | S9 | S10 | S11 | S12 | S13 | S14 | E1 | E2 | E3 | E4 | E5 | E6 | E7 | E8 | E9 | E10 |
| --- | --- | --- | --- | --- | --- | --- | --- | --- | --- | --- | --- | --- | --- | --- | --- | --- | --- | --- | --- | --- | --- | --- | --- | --- | --- | --- | --- | --- | --- |
| Age | 1.0 |  |  |  |  |  |  |  |  |  |  |  |  |  |  |  |  |  |  |  |  |  |  |  |  |  |  |  |  |
| Sex | .38 | 1.0 |  |  |  |  |  |  |  |  |  |  |  |  |  |  |  |  |  |  |  |  |  |  |  |  |  |  |  |
| H | -.23 | <u>-.01</u> | 1.0 |  |  |  |  |  |  |  |  |  |  |  |  |  |  |  |  |  |  |  |  |  |  |  |  |  |  |
| P | <u>-.14</u> | <u>-.11</u> | .47 | 1.0 |  |  |  |  |  |  |  |  |  |  |  |  |  |  |  |  |  |  |  |  |  |  |  |  |  |
| SA | <u>.11</u> | <u>.11</u> | <u>.06</u> | .35 | 1.0 |  |  |  |  |  |  |  |  |  |  |  |  |  |  |  |  |  |  |  |  |  |  |  |  |
| S1 | -.20 | <u>-.06</u> | .27 | .29 | <u>.04</u> | 1.0 |  |  |  |  |  |  |  |  |  |  |  |  |  |  |  |  |  |  |  |  |  |  |  |
| S2 | -.22 | <u>.03</u> | <u>.06</u> | <u>.19</u> | <u>.13</u> | .65 | 1.0 |  |  |  |  |  |  |  |  |  |  |  |  |  |  |  |  |  |  |  |  |  |  |
| S3 | -.19 | <u>.01</u> | <u>.13</u> | <u>.15</u> | <u>.11</u> | .72 | .71 | 1.0 |  |  |  |  |  |  |  |  |  |  |  |  |  |  |  |  |  |  |  |  |  |
| S4 | -.23 | <u>-.10</u> | <u>.15</u> | <u>.13</u> | <u>.07</u> | .39 | .56 | .51 | 1.0 |  |  |  |  |  |  |  |  |  |  |  |  |  |  |  |  |  |  |  |  |
| S5 | -.25 | <u>.00</u> | .23 | <u>.14</u> | <u>-.05</u> | .39 | .51 | .50 | .70 | 1.0 |  |  |  |  |  |  |  |  |  |  |  |  |  |  |  |  |  |  |  |
| S6 | -.22 | <u>.02</u> | .17 | <u>.17</u> | <u>.08</u> | .38 | .51 | .56 | .49 | .63 | 1.0 |  |  |  |  |  |  |  |  |  |  |  |  |  |  |  |  |  |  |
| S7 | -.27 | <u>.02</u> | <u>.14</u> | <u>.16</u> | <u>.00</u> | .42 | .39 | .57 | .50 | .53 | .48 | 1.0 |  |  |  |  |  |  |  |  |  |  |  |  |  |  |  |  |  |
| S8 | -.16 | <u>-.09</u> | <u>.13</u> | .18 | <u>-.06</u> | .40 | .46 | .53 | .37 | .27 | .33 | .33 | 1.0 |  |  |  |  |  |  |  |  |  |  |  |  |  |  |  |  |
| S9 | -.21 | <u>-.02</u> | .21 | .18 | <u>.06</u> | .55 | .66 | .54 | .63 | .64 | .59 | .54 | .42 | 1.0 |  |  |  |  |  |  |  |  |  |  |  |  |  |  |  |
| S10 | -.25 | <u>.03</u> | <u>.10</u> | .20 | <u>.09</u> | .40 | .62 | .51 | .55 | .64 | .55 | .62 | .28 | .74 | 1.0 |  |  |  |  |  |  |  |  |  |  |  |  |  |  |
| S11 | -.25 | <u>.01</u> | .28 | <u>.16</u> | <u>.01</u> | .60 | .58 | .65 | .43 | .49 | .58 | .46 | .52 | .51 | .46 | 1.0 |  |  |  |  |  |  |  |  |  |  |  |  |  |
| S12 | -.40 | <u>-.15</u> | <u>.13</u> | <u>.03</u> | <u>-.12</u> | .24 | <u>.13</u> | .21 | <u>-.01</u> | .17 | <u>.12</u> | <u>.12</u> | <u>.04</u> | <u>.07</u> | <u>.09</u> | .28 | 1.0 |  |  |  |  |  |  |  |  |  |  |  |  |
| S13 | <u>-.12</u> | <u>.09</u> | <u>.05</u> | <u>.14</u> | <u>.02</u> | .31 | .36 | .28 | .24 | .31 | .29 | .31 | .29 | .26 | .46 | .26 | <u>.14</u> | 1.0 |  |  |  |  |  |  |  |  |  |  |  |
| S14 | -.23 | <u>-.14</u> | .22 | .27 | <u>.00</u> | .58 | .59 | .61 | .52 | .53 | .46 | .41 | .56 | .53 | .48 | .60 | .35 | .30 | 1.0 |  |  |  |  |  |  |  |  |  |  |
| E1 | <u>.13</u> | -.20 | <u>.02</u> | <u>.06</u> | <u>.02</u> | <u>-.02</u> | <u>-.11</u> | <u>-.11</u> | <u>-.10</u> | <u>-.13</u> | <u>-.09</u> | -.26 | -.18 | <u>-.13</u> | -.24 | <u>-.12</u> | <u>.01</u> | -.16 | <u>-.02</u> | 1.0 |  |  |  |  |  |  |  |  |  |
| E2 | -.23 | -.26 | .18 | .24 | <u>.03</u> | .36 | .33 | .46 | .34 | .31 | .28 | .30 | .31 | .41 | .34 | .33 | .21 | .24 | .47 | <u>.16</u> | 1.0 |  |  |  |  |  |  |  |  |
| E3 | <u>.07</u> | <u>-.11</u> | <u>-.07</u> | <u>.06</u> | <u>.10</u> | <u>-.03</u> | <u>-.05</u> | <u>.06</u> | <u>-.06</u> | <u>-.03</u> | <u>.02</u> | <u>-.12</u> | <u>-.01</u> | <u>-.10</u> | <u>-.09</u> | <u>.07</u> | .15 | -.28 | <u>.08</u> | .33 | .17 | 1.0 |  |  |  |  |  |  |  |
| E4 | <u>-.13</u> | -.18 | <u>.14</u> | .15 | <u>-.12</u> | .23 | .28 | .24 | .20 | .31 | <u>.16</u> | <u>.06</u> | .32 | .22 | <u>.15</u> | .20 | <u>.14</u> | .24 | .36 | <u>.09</u> | .53 | <u>-.07</u> | 1.0 |  |  |  |  |  |  |
| E5 | <u>.11</u> | <u>-.03</u> | <u>.05</u> | <u>.01</u> | <u>.02</u> | -.28 | -.32 | -.45 | -.29 | -.36 | -.38 | -.50 | -.24 | -.26 | -.30 | -.39 | <u>-.08</u> | -.22 | -.21 | .24 | -.26 | <u>.05</u> | <u>-.16</u> | 1.0 |  |  |  |  |  |
| E6 | -.19 | -.22 | .21 | .21 | <u>-.08</u> | .29 | .25 | .26 | .37 | .37 | .32 | .18 | .21 | .27 | .24 | .18 | .19 | <u>.05</u> | .36 | .17 | .51 | .25 | .50 | <u>-.08</u> | 1.0 |  |  |  |  |
| E7 | <u>-.08</u> | -.50 | <u>.06</u> | <u>.06</u> | <u>.05</u> | <u>.04</u> | <u>.03</u> | <u>.09</u> | <u>.06</u> | <u>.00</u> | <u>.08</u> | <u>-.11</u> | <u>-.02</u> | -.15 | <u>-.11</u> | <u>.04</u> | <u>.13</u> | <u>-.11</u> | <u>.04</u> | .49 | .25 | .31 | .24 | <u>.11</u> | .28 | 1.0 |  |  |  |
| E8 | <u>-.07</u> | -.31 | <u>.09</u> | <u>.13</u> | <u>.00</u> | <u>.10</u> | <u>.00</u> | <u>.09</u> | <u>.15</u> | <u>.08</u> | <u>.03</u> | <u>-.10</u> | <u>-.11</u> | <u>-.07</u> | <u>-.01</u> | <u>.12</u> | <u>.07</u> | -.18 | <u>.04</u> | .35 | .20 | .35 | <u>.10</u> | <u>.14</u> | .41 | .60 | 1.0 |  |  |
| E9 | -.18 | -.19 | .19 | .28 | <u>-.01</u> | <u>.14</u> | <u>.13</u> | .20 | .24 | <u>.09</u> | <u>.01</u> | <u>.03</u> | <u>.14</u> | <u>.04</u> | <u>.06</u> | <u>.03</u> | <u>.12</u> | <u>-.02</u> | <u>.12</u> | .22 | .44 | .18 | .44 | <u>-.02</u> | .52 | .24 | .32 | 1.0 |  |
| E10 | <u>.06</u> | -.24 | <u>-.03</u> | <u>.09</u> | <u>.04</u> | <u>-.12</u> | -.16 | <u>-.09</u> | <u>-.09</u> | -.23 | -.10 | -.34 | -.19 | -.23 | -.20 | -.17 | -.21 | -.19 | -.19 | .22 | <u>-.01</u> | .34 | <u>.08</u> | .30 | .21 | .32 | .54 | .26 | 1.0 |

*Note.* H = daily number of hours treating COVID-19 patients; P = daily number of COVID-19 patients treated; SA = perceived safety provided by the protective equipment; S1-S14 = Perceived Stress Scale items; E1-E10 = Emotional Regulation Questionnaire items. The coding for the variable sex was 0 for females and 1 for males. Items S1-S14 and E1-E10 are treated as ordinal-categorical and the correlations between them are polychoric correlations. The remaining correlations including any of the rest of the variables are Pearson correlations. Non-significant correlations ( $p \geq .05$ ) appear underlined.

